# *ENOX1, CCDC122* AND *LACC1* ROLE IN PROGRESSION OF PROSTATE CANCER

**DOI:** 10.1101/2023.10.12.23296974

**Authors:** Timothy Ongaba

## Abstract

Prostate cancer (PCa) continues to trend among top 3 cancers that kill men over 20 years in the United Kingdom and worldwide despite extensive research and resources directed towards its treatment and prevention. In the application of a hallmark of survival mechanisms by the cancer, our study used mRNA seq data to identify genes that are increasingly mutated with progressing PCa from a cohort of 491 PCa patients. We found that *ENOX1, CCDC122* and *LACC1* deep deletion was positively associated with increasing age of diagnosis. Pathway analysis of enriched genes, following their deep deletion identified estrogen biosynthesis, KSRP signalling, omega 3 and 6 fatty acid biosynthesis and, Rap1 signalling as the top 5 enriched pathways. Previous individual and combined role of these genes in PCa progression was not fully established but thanks to this study, these pathways could be druggable targets in PCa patients with these gene deep deletions.

## Introduction

### Background to the investigation

Prostate cancer (PCa) is the most frequently diagnosed cancer in men in 132 countries with a 7.3% incidence worldwide. It is the second cause of cancer deaths in men in 48 of 185 sampled countries worldwide^1,2^. One in 8 men will be diagnosed with prostate cancer in the UK while one dies from the disease every 45 minutes. The number of PCa cases are projected to rise by 15% in the UK between 2023 to 2025 indicating an ever-increasing trend ^3^.

PCa starts *in-situ* within the prostate where it can, but not always, metastasize to the lymph nodes, bone, lung, liver and even as far as the brain when left untreated^4^. PCa can be termed localised when it remains within the prostate or metastatic when it spreads beyond the prostate. Localised PCa becomes high risk based on clinical staging, biochemical recurrence and a high Gleason score^5^.While still localised but more often when metastasised, PCa can progress from hormone sensitive to refractory phenotype with increasing resistance to treatment and decreasing survival rate from 30% over 5 to 30% over 3 years patient survival^6^.

When metastasised, the disease can be categorised as metastatic castration-sensitive prostate cancer (mCSPC) when clinical deprivation of androgens halts cancer advancement. The disease is termed castration-resistant prostate cancer(mCRPC) when cancer continues to progress even in the absence of androgens. The risk of fatality significantly increases when PCa becomes castration resistant^5^.

The incidence of PCa increases with age starting at 45, steeply rising and peaking around 75 to 79years. Patient survival is dependent on regular screening recommended to start at 45 years, early diagnosis and treatment intervention which might include chemotherapy, radiotherapy, surgical resection of tumours, immunotherapy or gland ablation to reduce the role of androgens in disease progression^3,7,8^.

Tumour Mutational Burden (TMB) of PCa, the somatic cell rate of mutation per mega base pair has been associated with low response to chemotherapy, high biochemical recurrence and, generally poor prognosis. These together with other cofounders such as poor immune cell infiltration into the tumour microenvironment, and androgen receptor status quickly lead to reduced survival of affected individuals^9–11^.

Among tumour mutations are gene deletions that can also appear as copy number alterations. Many of these, as deep deletions at chromosome 13q have been previously liked to poor prognosis in prostate cancer, high Gleason scores and biochemical recurrence^12^

Conversely, the high TMB also brings about high immunogenicity and, therefore, better response to immunotherapy^10,13^. This could, however apply to mutations in which the change in gene sequence produces a more antigenic protein product detectable by the affected individual. A deletion would lead to structural or functional alteration of the translated protein product.

Some of the most implicated genes in PCa are those involved in cell cycle progression, receptor signalling, proto-oncogenes, and or tumour suppressors. Among these, *UBE2C, PLK1, CDC20, BUB1, CDK1* and *HJURP* are top 6 linked to disease progression, metastasis and, poor prognosis^9,10^.

Ecto-NOX Disulfide-Thiol Exchanger 1(*ENOX1*), also known as cCNOX, CNOX, FLJ10094, and PIG38, is a copper-binding protein expressed in endothelial and other cells where it has NADH oxidase activity, promotes angiogenesis and is a potential druggable target ^14–16^. It is one of the genes on the 13q chromosome arm whose deletion is associated with poor prognosis, early biochemical recurrence and metastasis^12,17^

*ENOX1* has been negatively associated with prostate cancer, which is characterized by structural rearrangements, most frequently including translocations between androgen-dependent genes and other genes^18^. The Protein Atlas shows that *ENOX1* is expressed in cancer tissue, including prostate cancer^19,20^. *ENOX1* shares a 66% sequence identity with *ENOX2* of the same protein family with demonstrated contribution to gastric cancer cell growth and survival^21^. The exact role of *ENOX1* in prostate cancer is, however, not fully understood, and further research is needed to determine its functions and prognostic values^16^.

Coiled-Coil Domain Containing 122 (*CCDC122*) is a gene that is overexpressed in prostate cancer as compared to healthy prostate tissue^22,23^. There is generally limited literature linking the gene or its products to prostate cancer and this warrants further studies following its differential expression and or mutation during the progression of disease. Single nucleotide polymorphisms in the gene have previously been linked to development of leprosy in case controlled cohorts of Chinese and Brazilian populations^24,25^

The Laccase Domain-Containing 1(*LACC1*) gene is a gene that encodes an oxidoreductase that promotes fatty-acid oxidation, with concomitant inflammasome activation, mitochondrial and NADPH-oxidase-dependent reactive oxygen species production, and anti-bacterial responses in macrophages^22,26–29^.In relation to prostate cancer, a transcriptome analysis of differentially expressed genes in methotrexate-resistant prostate cancer found that *LACC1* was one of the genes that was differentially expressed^30^. A study aimed at identifying biomarkers for the management of human prostate cancer also found that *LACC1* was one of the genes that was differentially expressed in prostate cancer tissue compared to normal tissue^31^.Data from the TNM plot shows a reduced expression of the gene in prostate cancer compared to normal prostate tissue^23^. *LACC1* has also been linked to Leprosy in similar studies as *CCDC122*^24,25^.

## Materials and Methods

### Data retrieval from cbioportal

Using mRNA Seq data from the Prostate adenocarcinoma (TCGA Firehose Legacy) in cbioportal (https://www.cbioportal.org/), we examined quartiles of increasing age at time of diagnosis to identify what genes are incrementally altered with increasing age of diagnosis^32^.

### Genomic alteration analysis of *ENOX1, CCDC122* and *LACC1*

Of 491 samples/patients that had mRNA expression z-scores relative to all samples (log mRNA Seq V2 RSEM) we used the onco query language “*ENOX1*: HOMDEL, *CCDC122*: HOMDEL, *LACC1*: HOMDEL” as the user defined list, the mRNA expression z-score relative to all samples, in addition to putative copy number alteration from GISTIC and mutations were queried from the study samples with mutation and Copy Number Alteration (CNA) data.

The z-score threshold was left at ±2.0 All other parameters were left as default.

#### Effect of *ENOX1, CCDC122* and *LACC1* in PCa

We examined mRNA seq data Z-score relative to all genes for patients that had deep deletions for each of the three genes to compare their individual and combined effect on the survival of PCa patients as overall survival.

#### Relationship analysis for ENOX1, CCDC122 and LACC1

Using data from String database (available at https://string-db.org/) the three protein names were searched for under the “Multiple proteins” tab for “*Homo sapiens*” as the organism^33^.

### Pathway analysis using Enrichr and Reactome

To examine what genes are upregulated following the deep deletion of the three genes, we found patient samples’ mRNA expression (RNA Seq V2 RSEM) (491 samples) data that had at least one deep deletion of *ENOX1, CCDC122* and *LACC1* (altered group of 81) and those with deep deletions of all three genes(unaltered).

As our interest was pathway analysis of genes implicated with deep deletion of all 3 genes (*ENOX1, CCDC122* and *LACC1*), we selected for the “unaltered” gene category list with statistically significant expression (95%CI) and ordered these by descending log2 ratio of the z-score absolute values then by ascending p-value. All 432 genes were used an input for pathway analysis in Enrichr (https://maayanlab.cloud/Enrichr/)and later Reactome (https://reactome.org/) for cross reference ^34,35^. The gene names were pasted in both analysis webpages and all parameters were left as default in the analysis tab.

Individual gene deep deletion effect on physiological pathways was not considered for analysis as patient data showed a multigene effect compounding disease status.

In Enricher, the most up-to-date manually curated pathway reference database and the highest combined score for the top 4 pathways identified were chosen as the best reference database for pathway analysis of the set of 432 genes.

### Statistical analysis

All statistical analysis was done within cbioportal.

## Results

### Data retrieval /overview of Prostate Adenocarcinoma study (TCGA, Firehose Legacy)

To examine for genes increasingly mutated with progressing disease, we chose a cohort of PCa patients with a wide range of age at time of diagnosis and Gleason primary and secondary patters. Our chosen patient cohort with mRNA seq data for 498 patient samples suited well for downstream analyses for PCa genomic analysis. (Figure1).

**Figure 1.**
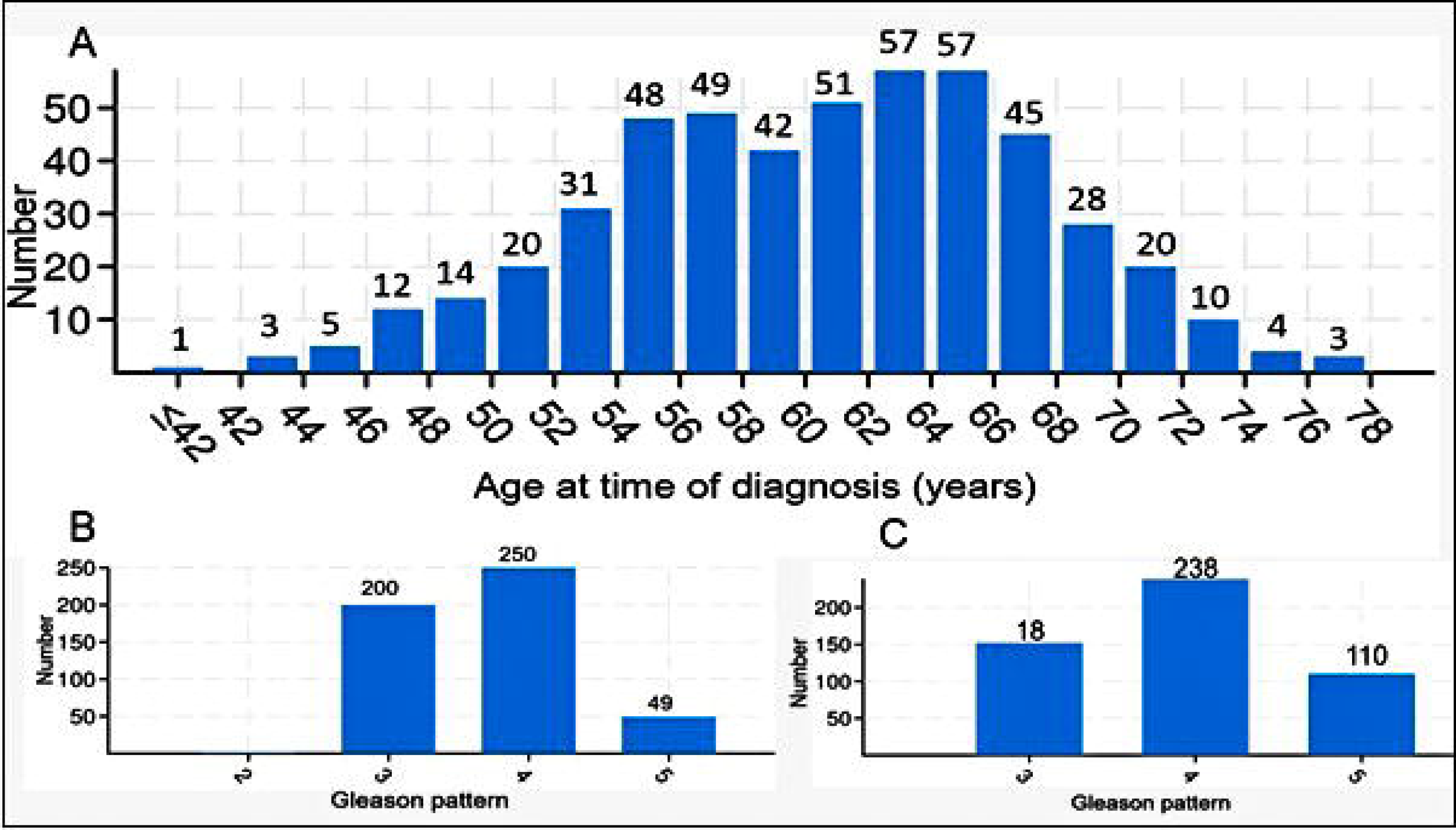
Wide scope of age of diagnosis and Gleason scores. A shows age class boundaries of patients diagnosed with PCa. B represents different primary whereas C shows the secondary Gleason patters making up different Gleason scores of PCa in patients.

#### Differential gene mutation with increasing age

Increasing age quartiles of PCa diagnosed patients showed differential genomic alterations. Three of the genomically altered genes; *ENOX1, CCDC122* and *LACC1* had increasing gene deep deletions with increasing age at time of diagnosis with 0.0883, 0.146 and 0.146 p-values respectively. (Figure 2)

**Figure 2.**
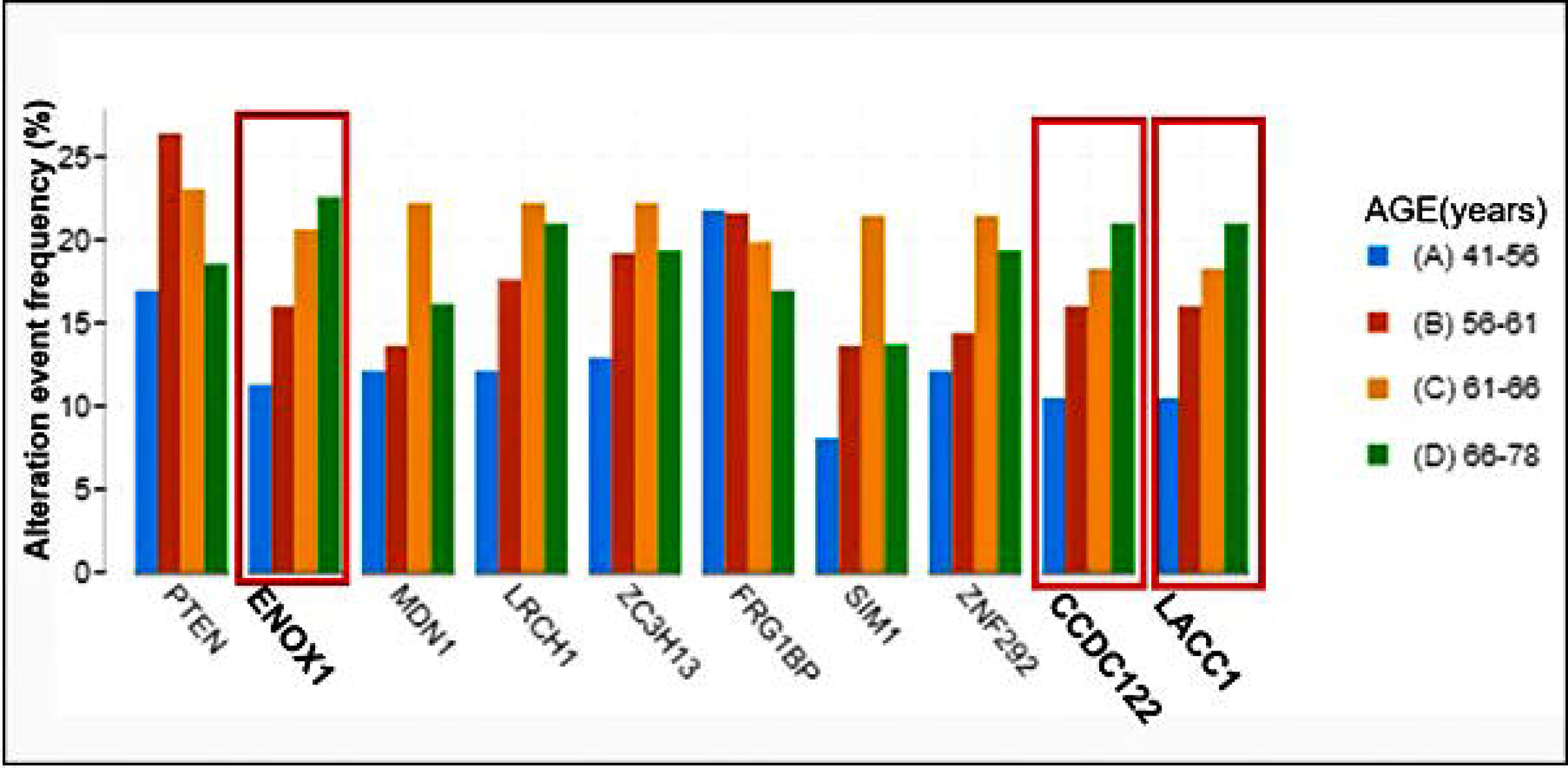
Genomically altered genes with increasing age quartiles. *ENOX1, CCDC122* and *LACC1* are increasingly altered with increasing age of diagnosis.

All three genes were located on the 13q14.11 cytogenetic band having a deep deletion mutation.

### Genomic alteration analysis of *ENOX1, CCDC122* and *LACC1*

Sixteen percent (16%) of the 491 patient cohort had deep deletions of all three genes. All the three gene deep deletions had a co-occurrence tendency with p-values all less than 0.001 (95% CI) (Table1).

**Table 1.** *ENOX1, CCDC122* and *LACC1* are concurrently deleted in in 491 patient samples.

| A | B | Neither | A Not B | B Not A | Both | Log2 Odds Ratio | p-Value | q-Value | Tendency |
| --- | --- | --- | --- | --- | --- | --- | --- | --- | --- |
| *ENOX1*:<br>HOMDEL | *CCDC122*:<br>HOMDEL | 412 | 0 | 0 | 79 | >3 | <0.001 | <0.001 | Co-occurrence |
| *CCDC122*:<br>HOMDEL | *LACC1*:<br>HOMDEL | 410 | 2 | 0 | 79 | >3 | <0.001 | <0.001 | Co-occurrence |
| *ENOX1*:<br>HOMDEL | *LACC1*:<br>HOMDEL | 410 | 2 | 0 | 79 | >3 | <0.001 | <0.001 | Co-occurrence |

Analysis of any mutational data on the 3 genes irrespective of expression profile showed no patient had any mutation in the protein product of any of the (Supplementary figure 1)

#### Effect of *ENOX1, CCDC122* and *LACC1* deep deletion in PCa

There was no significant difference in overall survival between patient who had a deep deletion and those who had an amplification of all three genes on a Kaplan-Meier plot (Supplementary figure 2)

#### Relationship analysis for ENOX1, CCDC122 and LACC1

CCDC122 and LACC1 proteins were found to be co-expressed with in addition to text mining results. ENOX1, had no shown relationship with either gene (Figure 3).

**Figure 3.**
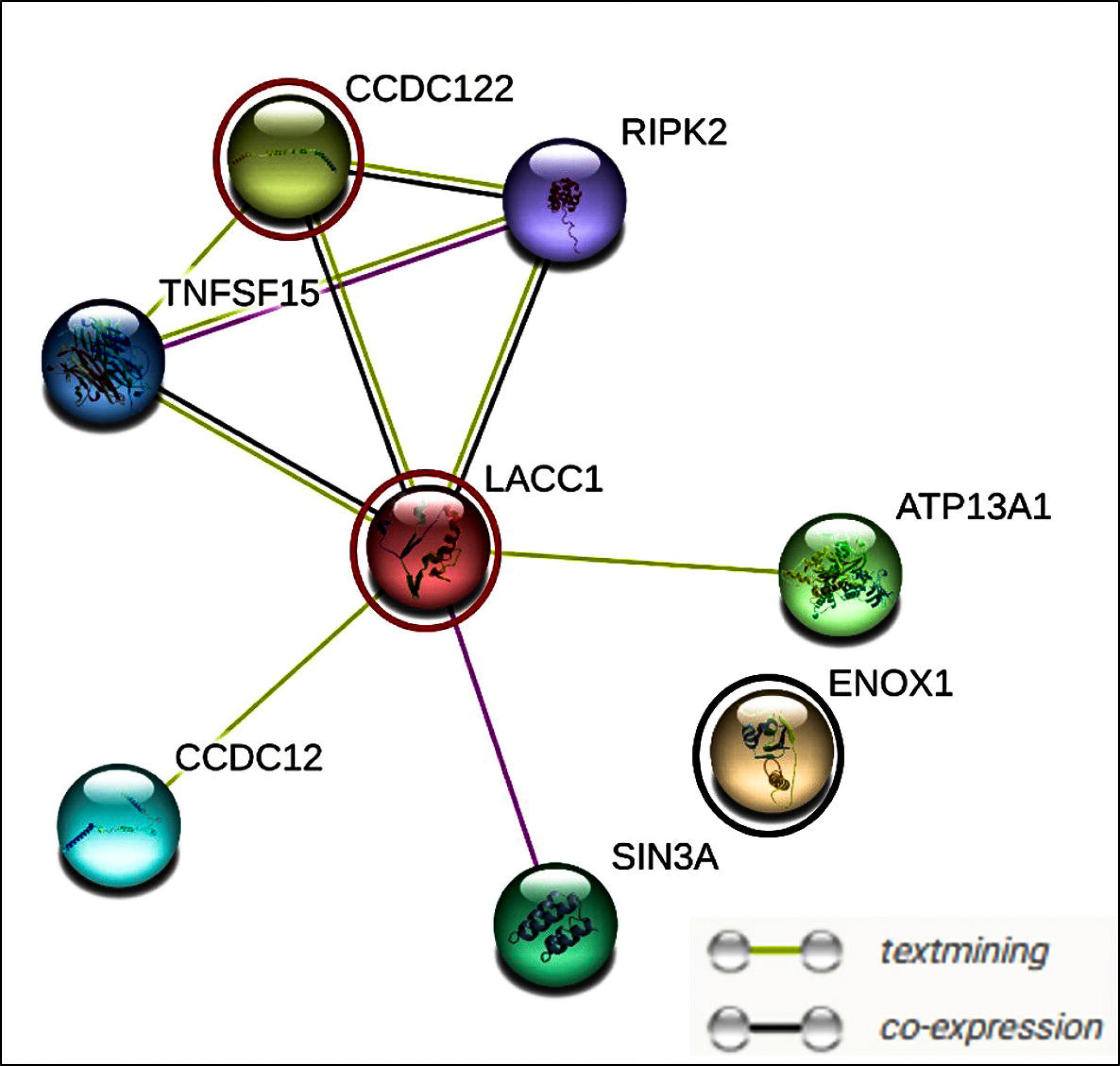
*ENOX1* has not protein relationship with, *LACC1* and *CCDC122* which were liked with evidence of co-expression and text mining The three putative interactions were circled in red (*LACC1* and *CCDC122*) and black(*ENOX1*).

### Pathway analysis using Enrichr and Reactome

Reactome2022 was the database with the highest combined scores and most up-to-date in Enrichr pathway analysis. The individual gene deep deletion implicated pathways were identical between *LACC1* and *CCDC122* with *ENOX1* having a more alike effect to the combined effect of all three gene deep deletions.

The top 5 identified pathways ranking first by descending combined score then by ascending p-value for all three genes’ deep deletion were estrogen biosynthesis, KH-type splicing regulatory protein(KSRP), alpha linoleic and linolelic acid metabolism, and Rap1 signalling with 114.62,110.08,82.59,67.35,56.94(2dp) combine scores respectively (Table 2).

**Table 2.** Top five upregulated genes were from estrogen biosynthesis, KSRP activity, omega 3 and, 6 and, Rap1 signalling pathway.

| Index | Name/Pathway Identifier | P-value | Odds Ratio | Combined score | Genes overlap |
| --- | --- | --- | --- | --- | --- |
| 1 | Estrogen Biosynthesis<br>R-HSA-193144 | **0.006562** | 22.8030303 | **114.6185** | *AKR1B15;CYP19A1* |
| 2 | KSRP (KHSRP) Binds and Destabilizes mRNA<br>R-HSA-450604 | **4.05E-04** | 14.09187534 | **110.0775** | *DIS3;EXOSC8;PARN;YWHAZ* |
| 3 | Alpha-Linolenic (Omega3) And Linoleic (Omega6) Acid Metabolism<br>R-HSA-2046104 | **0.00242** | 13.70957944 | **82.58704** | *ELOVL1;MUSK;SYCP2* |
| 4 | Linoleic Acid (LA) Metabolism<br>R-HSA-2046105 | **0.011904** | 15.2004662 | **67.3514** | *ELOVL1;MUSK* |
| 5 | Rap1 Signalling<br>R-HSA-392517 | **0.004516** | 10.5442128 | **56.93941** | *RAP1B;RAP1A;YWHAZ* |

Results from Reactome corroborated the same pathways as identified by Enrichr displaying genes that overlapped with those from known pathways (False Discovery Rate (FDR)<0.001). The top two of these with respective overlapping genes were highlighted (Figure 4). 184 genes were not mapped to any pathway by Reactome.

**Figure 4.**
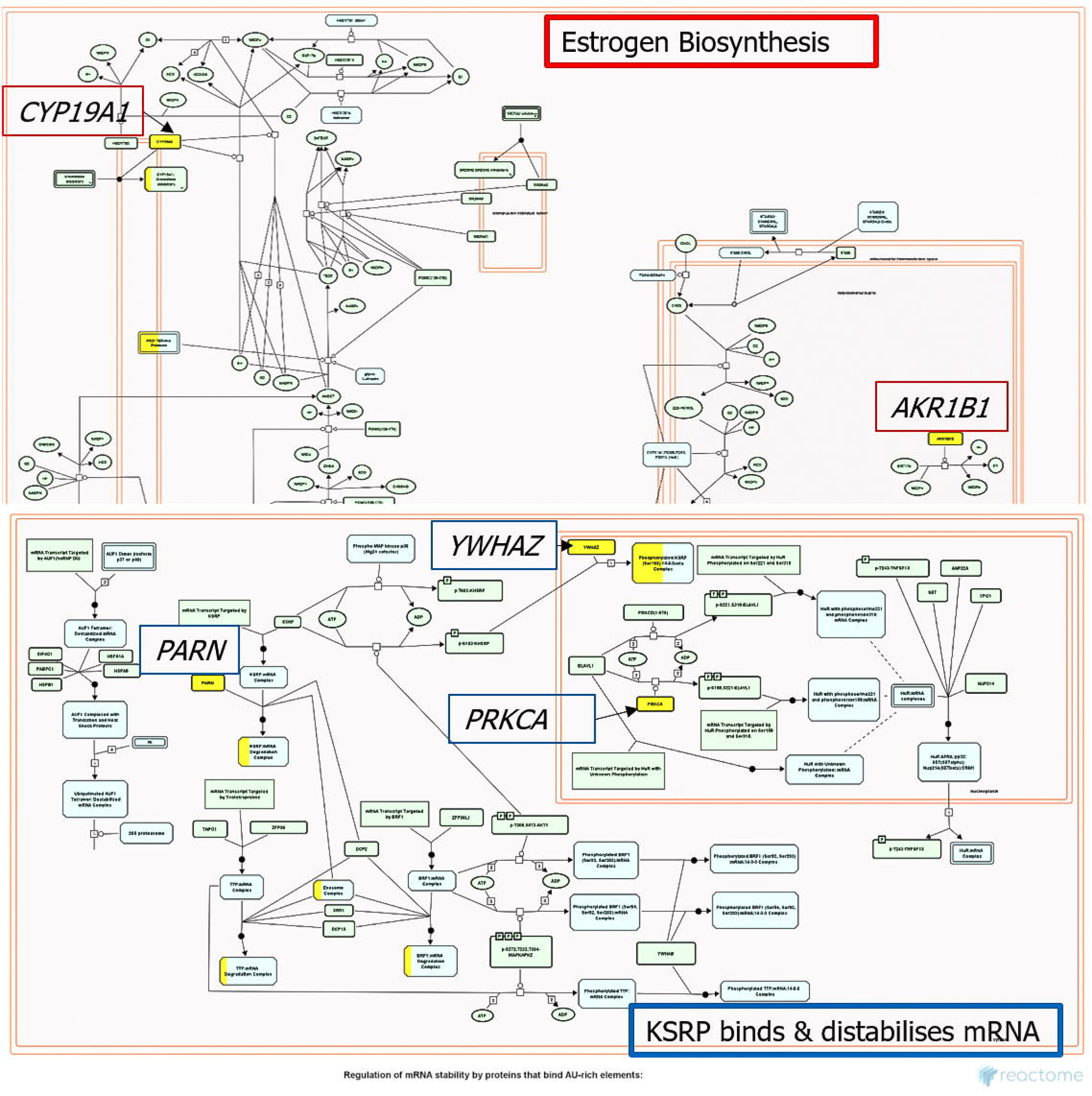
Gene’s overlap (coloured yellow) between query genes mapped to known pathways by Reactome. Top is the estrogen biosynthesis pathway and bottom is the KSRP binding and destabilisation of mRNA pathway.

## Discussion

Our study aimed to identify some of the most prevalent genomic alterations and or differentially expressed genes throughout the progression of disease. As age at diagnosis is often positively correlated and had previously been found to be linked with disease progression and poor prognosis, we examined quartiles of ages of patients diagnosed with prostate cancer to identify those genes whose alteration frequency was increasing with age and Gleason score(Figure1)^36,37^.

Whereas the development and progression of prostate cancer is not pivotal on a single or only genomic mutations, we observed roles such as structural alterations and fusions attributed to TMPRSS2 gene and other chromosome 13q gene deletions among a hallmark of other factors PCa employs to develop, evade and progress in an individual. *ENOX1*, one of our identified genes is found on the same chromosome arm and we were able to identify its combined role with *CCDC122* and *LACC1* within the same cytogenetic band of 13q14.11 in progression of PCa as coexpression^38,39^.

All the top 5 pathways implicated by upregulated genes following deep deletion of *ENOX1, LACC1* and, *CCDC122* have been previously linked to cancer progression and give insight on the beneficial effect of the three gene deletion to prostate cancer cell survival and disease progression. The enriched pathways as a result of all three gene deletions were identical to those enriched by *ENOX1* single gene deletion alone whereas single gene deletion of *LACC1* and *CCDC122* had the same not identical effect to the pathways enriched by all three gene deletions (Supplementary figure 3). This suggests that the overall observed enriched pathways were more as a result of single gene deletion of *ENOX1* that either or both of *LACC1* and *CCDC122*.

Estrogen has been shown to play a role in the development and progression of prostate cancer through its receptor activation on PCa cells that promotes cell proliferation and survival^40–42^. Estrogen can stimulate the production of growth factors and cytokines that promote angiogenesis and tumour growth^40,41,43^. Estrogen can inhibit the androgen receptor (AR) signalling pathway, and this is important for prostate cancer growth and survival^40,43^.

KSRP (KH-type splicing regulatory protein) is an RNA-binding protein that has been shown to promote tumour growth and metastasis in non-small cell lung, breast, ovarian and, prostate cancer^44^. A study on castration-resistant prostate cancer showed that KSRP regulates type III interferon expression post-transcriptionally, which is an important axis in the progression of prostate cancer especially in advanced stage^45^. KSRP is also known to regulate gene expression on various levels, including gene transcription, mRNA decay, and microRNA precursor maturation^44^. In colorectal cancer, enhanced expression of KSRP was found in tumour tissue and was associated with worse overall survival. KSRP seemed to drive epithelial cell proliferation in primary and metastatic cells through control of cell cycle progression and promoted angiogenesis by enhancing vascular endothelial growth factor^46^.

Omega-3 and omega-6 fatty acids play a crucial role in the development and progression of prostate cancer. High amounts of omega-6 fatty acids have been linked with an increased risk of prostate cancer, whereas omega-3 fatty acids have been shown to inhibit prostate cancer growth^47^. A study on athymic mouse xenograft model simulating radical prostatectomy showed that prostate tumours can be modulated by the manipulation of omega-6:omega-3 ratios through diet, and the omega-3 fatty acid stearidonic acid (SDA) (precursor of eicosapentaenoic acid (EPA)) can inhibit prostate tumour growth and recurrence^48^. However, a study found that high intake of omega-3 fats in the blood is linked to an increased risk of prostate cancer^49^. Another study showed that there were no significant associations between specific omega-3 or omega-6 PUFA intakes and overall prostate cancer risk^50^. Our current findings show a beneficial role of these two essential acids in PCa.

Ras-proximate-1 (Rap1) signalling has been shown to play a role in prostate cancer metastasis and invasion^51–53^. Specifically, Rap1 promotes tumour progression by disrupting E-cadherin-mediated cell adhesion, leading to a reduction in E-cadherin^54^. Activation of Rap1 has also been shown to increase prostate cancer cell migration and invasion through the α4, β3, and αvβ3 integrins^53^. Additionally, a study found that miR-203 down-regulates Rap1A and subsequently suppresses cell proliferation, adhesion, and invasion in prostate cancer^53^. These studies summatively suggest Rap1 contributes to PCa metastasis and, invasion in patients through several pathways

Whereas our initial supposition was that *ENOX1, LACC1* and *CCDC122* were increasingly expressed to benefit PCa, our findings show that their deletion instead enriches genes that achieve the same outcome of PCa cell growth and survival.

The novelty in our findings is the link between the three identified genes whose combined deletions enriches genes in in PCa cell benefiting pathways for disease cell growth and survival. The three gene protein products, however, have positive physiological body functions so it is putative that on deletion, redundant functionality from similar proteins whose deletion would be detrimental to PCa cells is employed as a substitute.

For a wet lab proof of concept, future work might involve use of PCa cell lines (RWPE-1, LNCaP, PC3, DU145) demonstrating different stage progression or stem cells of the disease and screening for the three genes using fluorescent *in-situ* hybridisation technique under the expectation of decreasing fluorescence with advancing disease.

As a therapeutic target for PCa and personalised therapy, judicial targeting of rate limiting steps/enzymes in the identified pathways could be explored as a treatment option should the side effect not outweigh the clinical benefit of the intervention.

## Data Availability

https://www.cbioportal.org/study/summary?id=prad_tcga

## Acknowledgments

Special appreciation to Dr. Amanda Coutts (P.H.d) and the School of Science and Technology for the guidance and resources used in executing this work.

