## SupplementaryFigures for "*ENOX1, CCDC122* AND *LACC1* ROLE IN PROGRESSION OF PROSTATE CANCER"

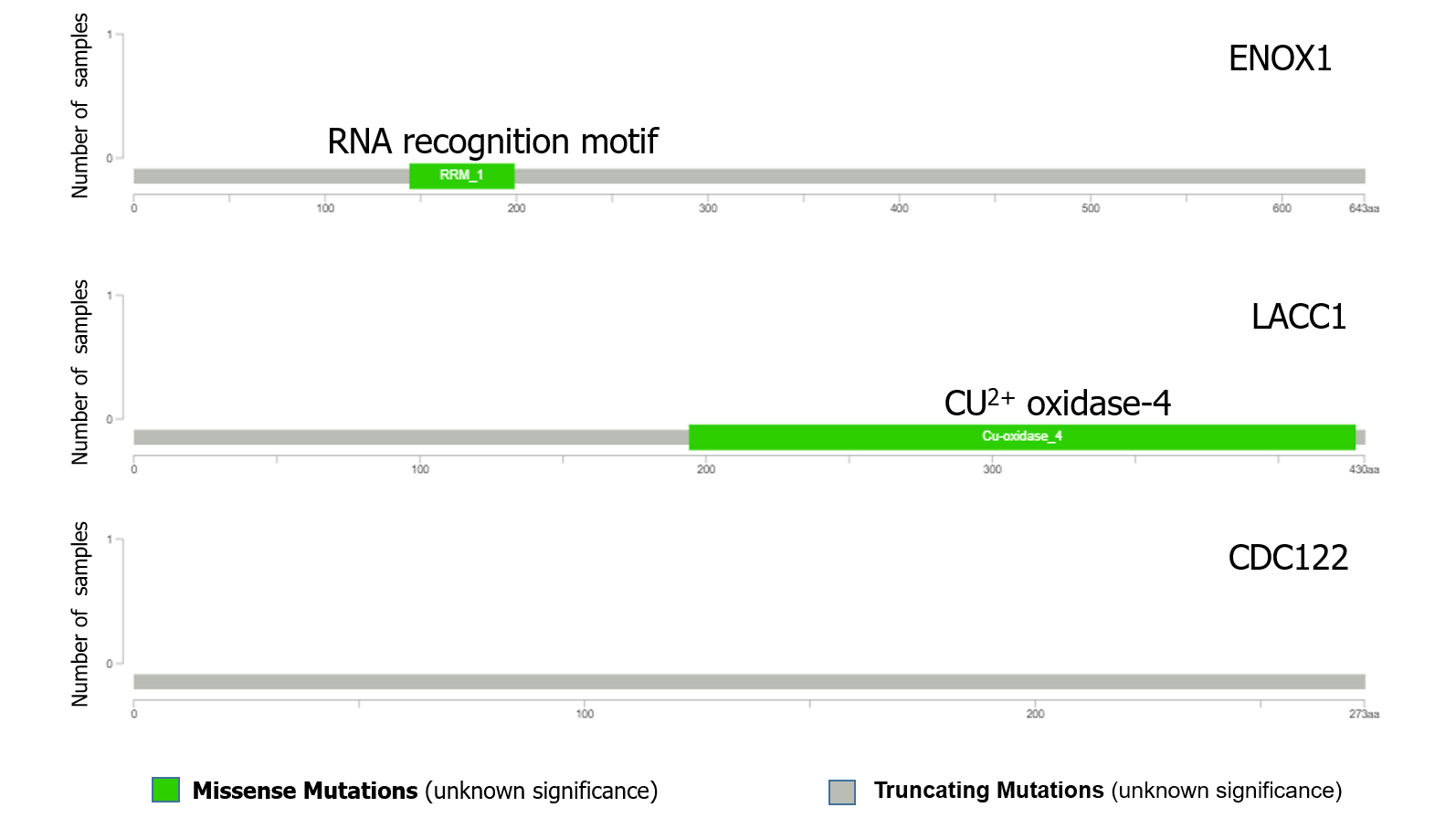

**Supplementary Figure 1|** Lollipop plots of ENOX1, LACC1 and CCDC122. There was no patient sample recorded for any amino acid mutations un any of the functional motifs.

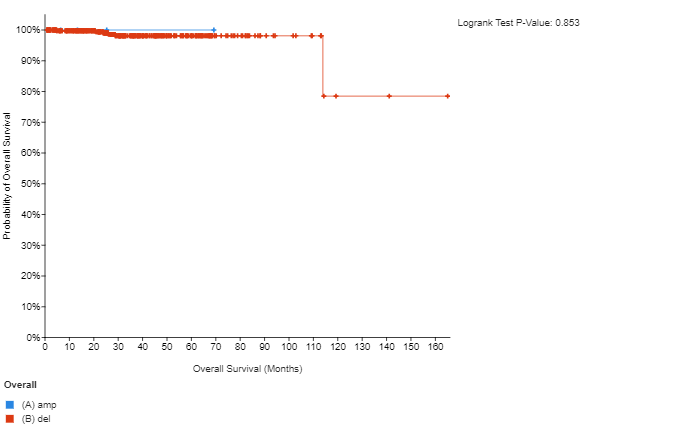

**Supplementary Figure 2**| Kaplan-Meier plot of patients with *ENOX1, LACC1* and *CCDC122* amplification versus those with deep deletion of all three genes. Deep deletion of *ENOX1, LACC1* and *CCDC122* even without statistical significance tend to be negatively associated with poor survival rate.

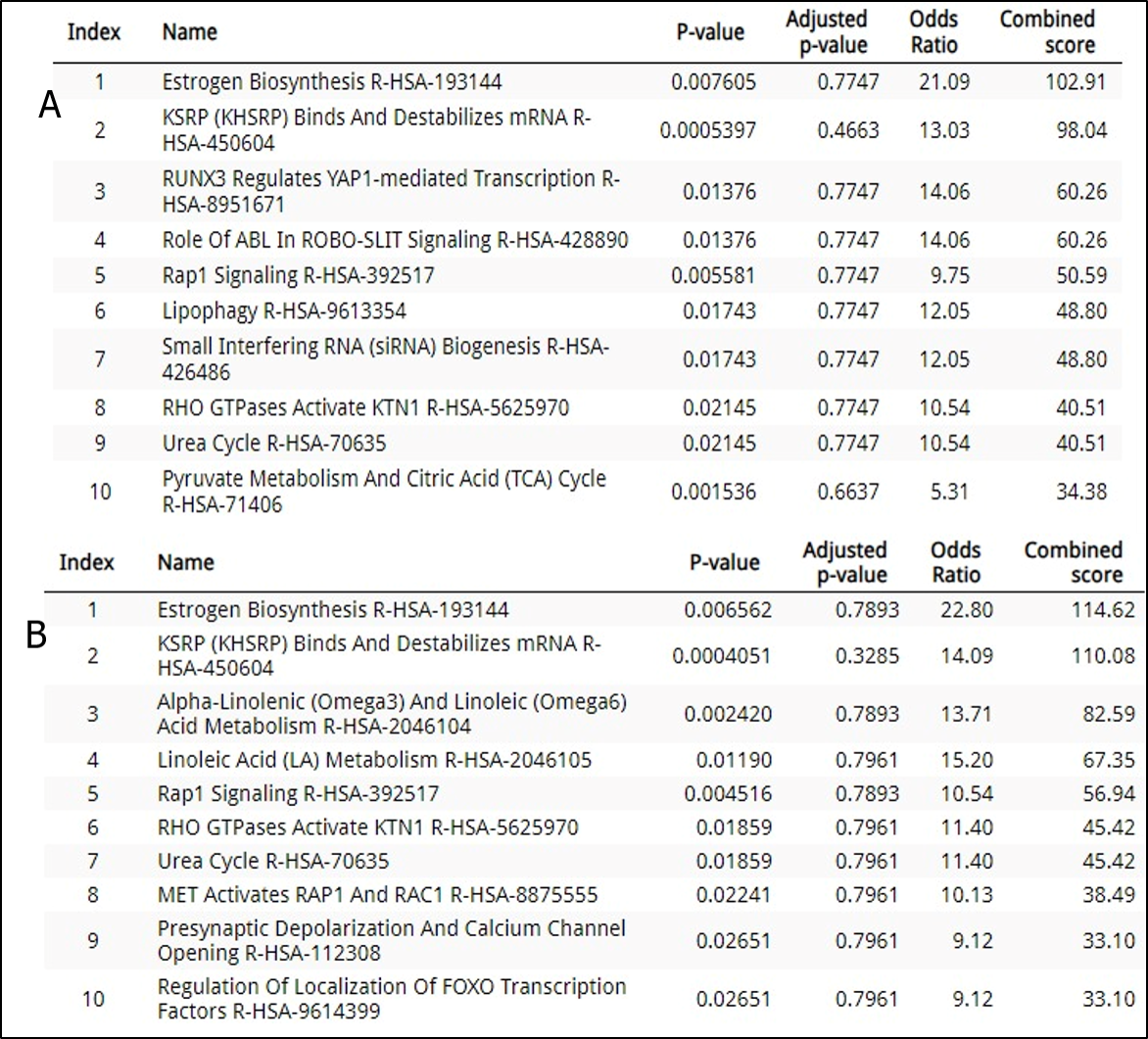

**Supplementary figure 3**| Pathways implicated by individual deep deletion of *LACC1*, *CCDC122* and *ENOX1.*A represents pathways from genes upregulated by deep deletion of *LACC1*, *CCDC122* individually. B shows pathways from genes upregulated by deep deletion of *ENOX1.*
